# Predicting epileptic seizures using nonnegative matrix factorization

**DOI:** 10.1101/19000430

**Authors:** Olivera Stojanović, Gordon Pipa

## Abstract

This paper presents a procedure for the patient-specific prediction of epileptic seizures. To this end, a combination of nonnegative matrix factorization (NMF) and smooth basis functions with robust regression is applied to power spectra of intracranial electroencephalographic (iEEG) signals. The resulting time and frequency components capture the dominant information from power spectra, while removing outliers and noise. This makes it possible to detect structure in preictal states, which is used for classification. Linear support vector machines (SVM) with L1 regularization are used to select and weigh the contributions from different number of not equally informative channels among patients. Due to class imbalance in data, synthetic minority over-sampling technique (SMOTE) is applied. The resulting method yields a computationally and conceptually simple, interpretable model of EEG signals of preictal and interictal states, which shows a good performance for the task of seizure prediction.

## Introduction

The ability to predict epileptic seizures provides an opportunity to intervene in order to attenuate their effects, or if possible prevent them. In this study we focus on EEG manifestations of seizures, which are characterized by sudden hypersynchronization of neurons and last from seconds to minutes. [1] Recently published studies on seizure prediction use a wide variety of approaches, from time series analysis (e.g. phase synchronization [2]) and spectral features of EEG signals [3] to physiological models of neural activity (e.g. neural mass models [4]). We focus on spectral measures of EEG signals since they have been successfully used as features for seizure prediction, and are easily interpretable. [3, 5, 6]

In the field of seizure prediction there are certain conceptional, computational and data-related challenges. First, using a large number of features for prediction makes it difficult to interpret their individual contribution. [6] Secondly, the algorithms for seizure prediction in a clinical setting need to be computationally efficient. Due to hardware constraints, this applies to closed-loop EEG devices for seizure prediction and intervention in particular, which have been a recent focus in the field. [5–7] Finally, data encountered in the field of seizure prediction can be high dimensional and heterogeneous (e.g. recorded using many different channels and types of measurements in addition to EEG, like ECG, EOG etc), yet suffer from class imbalance (patients spend more time in interictal than in preictal states) and limited in the number of labeled samples. This is particularly challenging for the design of a patient-specific model.

In this study we address these issues by developing an easy-to-use, computationally efficient method for patient-specific seizure prediction. In order to achieve that, we extract a small set of interpretable features from power spectra that distinguish a baseline (interictal) EEG activity from a state leading up to a seizure (preictal state). Interictal states are regular brain activity between seizures, which can sometimes be interrupted with interictal spiking. [1, 8] Since seizures are characterized by strong synchronization, they are very prominent in power spectra of EEG signals. Although preictal states are not clearly visible in raw EEG signals, multiple studies confirmed the presence of distinct preictal states using spectral [3, 9, 10], as well as information measures. [11–13] For a detailed discussion, see [5] and [6].

Although power spectra capture relevant changes in frequency over time, they can be very noisy and contain outliers. We thus use nonnegative matrix factorization (NMF) [14, 15] to decompose power spectra into dominant time and frequency components, which are later used for seizure prediction.

To mitigate class imbalance, we employ synthetic minority over-sampling technique (SMOTE) [16], together with linear SVM with L1 regularization, to assign weights for contributions from each individual channel and eliminate uninformative channels. The developed method is computationally inexpensive and produces good results while providing insights into the structure of preictal states.

## Materials and Methods^1^

### Data preparation

The data consists of heterogeneous EEG recordings of six patients (two females; median age: 33) [Tab.1] and form a part of the bigger Freiburg EPILEPSIAE database. [17] Recordings were made at the University Medical Center Freiburg, over the course of several days (three to nine), between 2003 and 2009. The sampling frequency varies between 256Hz and 1024Hz. The electrodes that are used in the recordings include intracranial (depth, strip and grid) and surface electrodes, together with special electrodes (e.g. ECG, EMG and EOG), whose number varies between 31 and 122, depending on the diagnosis. In order to investigate preictal states thoroughly, only intracranial EEG recordings are used.

Since the ability to predict a seizure five minutes before its onset can be useful for patients with uncontrolled epilepsy [18], we focus on five minute intervals of preictal and interictal states. In the case of a preictal state, an interval of five minutes leading up to a seizure is extracted. Since preictal states directly precede seizures, seizure prediction can be realized by classification between preictal and interictal states.

In the case of an interictal state five minutes intervals are extracted, which are at least 11 minutes before or after any other seizure. We refer to these intervals of extracted signals as individual measurement periods. The data are filtered with the Parks-McClellan optimal equiripple finite impulse response filter to remove 50Hz line noise.

The dataset is separated into training (70%) and validation set (30%) during a 100-fold cross-validation procedure.

**Table 1.** Detailed information about patients the from EPILEPSIAE database. [17]

| Patient's number | age | sex | number of channels | sampling frequency (Hz) | number of preictal intervals | number of interictal intervals |
| --- | --- | --- | --- | --- | --- | --- |
| 1 | 34 | male | 48 | 256 | 23 | 88 |
| 2 | 37 | female | 26 | 512 | 7 | 44 |
| 3 | 18 | male | 94 | 1024 | 9 | 80 |
| 4 | 42 | male | 38 | 1024 | 7 | 110 |
| 5 | 15 | male | 91 | 256 | 17 | 9 |
| 6 | 32 | female | 68 | 1024 | 7 | 19 |

### Deriving time and frequency components

To identify stereotypical behavior between and ahead of seizures, spectrograms of each channel [Fig.1] are obtained using the multitaper method [19] with time windows of 10 seconds (which is calculated by using 50% overlap of a 20 seconds window). To correct for baseline activity across frequencies in interictal states, relative power is calculated by dividing spectrograms of each channel by the respective average interictal spectrogram.

Due to the clinical setting and patients’ diagnoses, the sampling frequency varies among different patients. As a result, the highest frequency in the spectrograms varies between 128Hz and 513Hz. However, this difference is unproblematic due to the fact that we develop patient-specific models. After obtaining spectrograms of every individual measurement period for every channel, they are visually inspected, and in the case of anomalies, excluded from the training data. In addition to that, if either the beginning or end of an individual measurement period is corrupted by e.g. seizure leakage (an onset of a seizure before a misplaced onset label due to hand-labeling of the raw iEEG signal) or electrode detachment, then the corrupted data is replaced by a corresponding amount of data from the opposite end of the measurement interval. In that way, the length of spectrograms is preserved.

### Time-frequency decomposition

To examine changes in power spectra, spectrograms of each channel and each individual measurement period are decomposed into a time and a frequency component using nonnegative matrix factorization. Originally proposed under the name “positive matrix factorization”, it is a variant of factor analysis [14], which was first used on environmental data [20] and later popularized in the application to face recognition under the current name. [15] For both tasks, NMF is successful in learning interpretable parts-based representation (e.g. concentrations of elements, as in [20] or parts of faces, as in [15]) and shown to perform better than independent component analysis, principal component analysis or vector quantization. [21–23] In the field of seizure prediction, NMF has been used to develop a method for automatic localization of epileptic spikes in children with infantile spasms [24] and for automatically detection and localization of interictal discharges. [25]

Nonnegative matrix factorization decomposes a nonnegative matrix *V* into two nonnegative low-rank matrices *W* and *H* [15]:

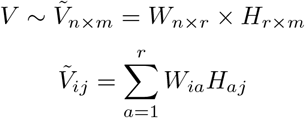

The outer product 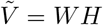 can be interpreted as a low rank parts-based approximation of the data in *V*. [15] We decide on a factorization of rank *r* = 1 to get the most constrained model with two vectors, one of which represents temporal evolution (time component *H*) and one of which represents distribution of frequencies (frequency component *W*). [Fig.2]

To lessen the influence of outliers and to remove noise in the NMF components, they are modeled with smooth basis functions using robust regression. The time component is modeled by a polynomial of second order, while the frequency component is modeled by nonlinearly logarithmically spaced B-splines of sixth order to consider the frequency resolution which decreases in higher frequencies. [Fig.2] By modeling each component with smooth basis functions, the most relevant information is preserved in both domains, while noise is removed.

By calculating the outer product of modeled NMF components as shown in figure 3, time-frequency models can be reconstructed. They capture the most important information while leaving out the noise and thus provide simplified intermediate representation of the data, which can be visually compared to the corresponding spectrograms (see S1 Figure in the appendix). The coefficients of the modeled time and frequency components therefore convey relevant information about structure of both states.

**Figure 1.**
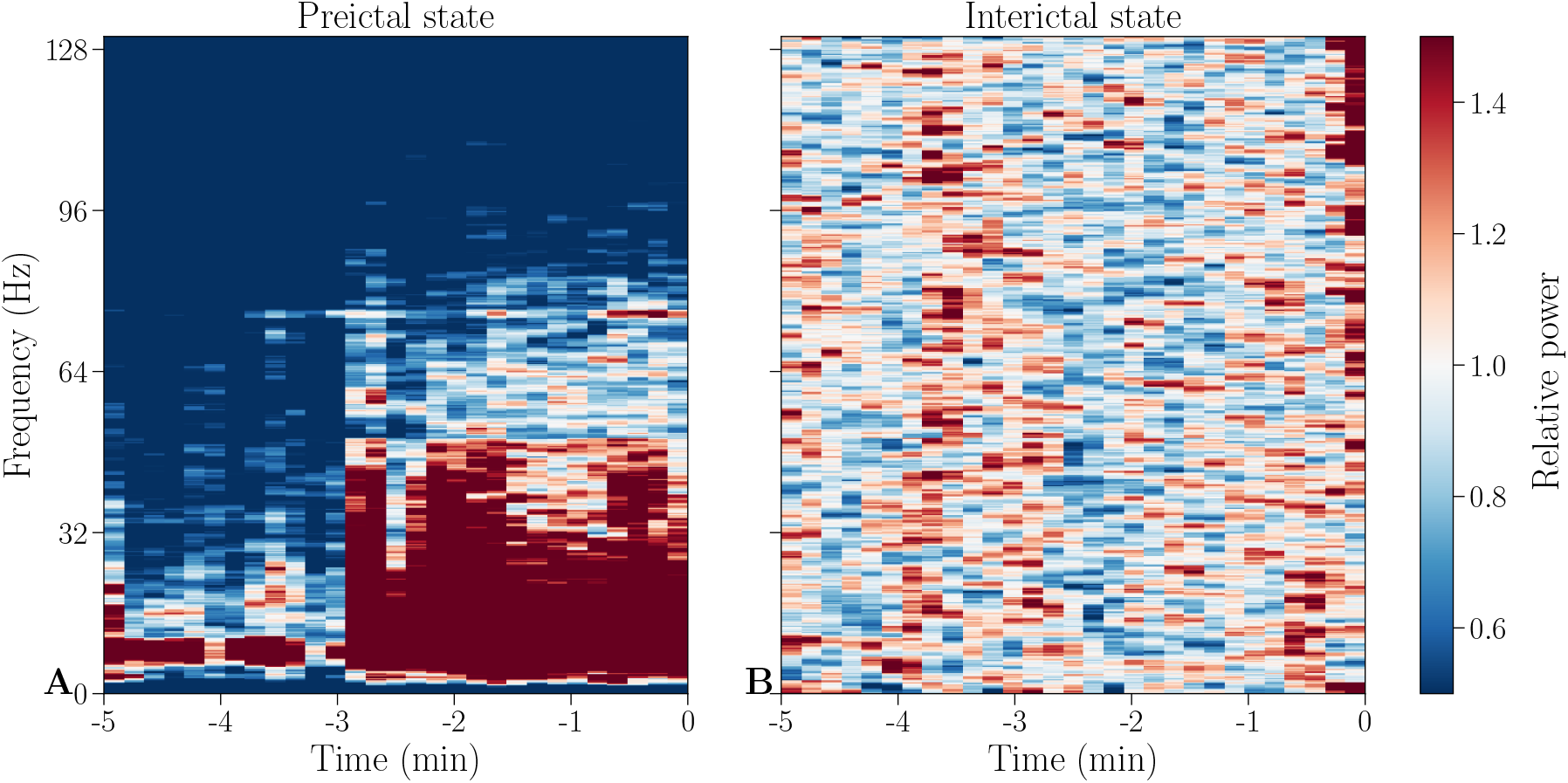
Example spectrograms of interictal and preictal states. Baseline corrected spectrograms of a preictal (**A**) and an interictal (**B**) individual measurement period of channel HR1 from patient 1. The preictal spectrogram shows an onset of a strong low frequency oscillation, while the interictal spectrogram is similar to the baseline activity. This channel and individual measurement period will be used throughout the paper for illustrative purposes, if not stated otherwise.

**Figure 2.**
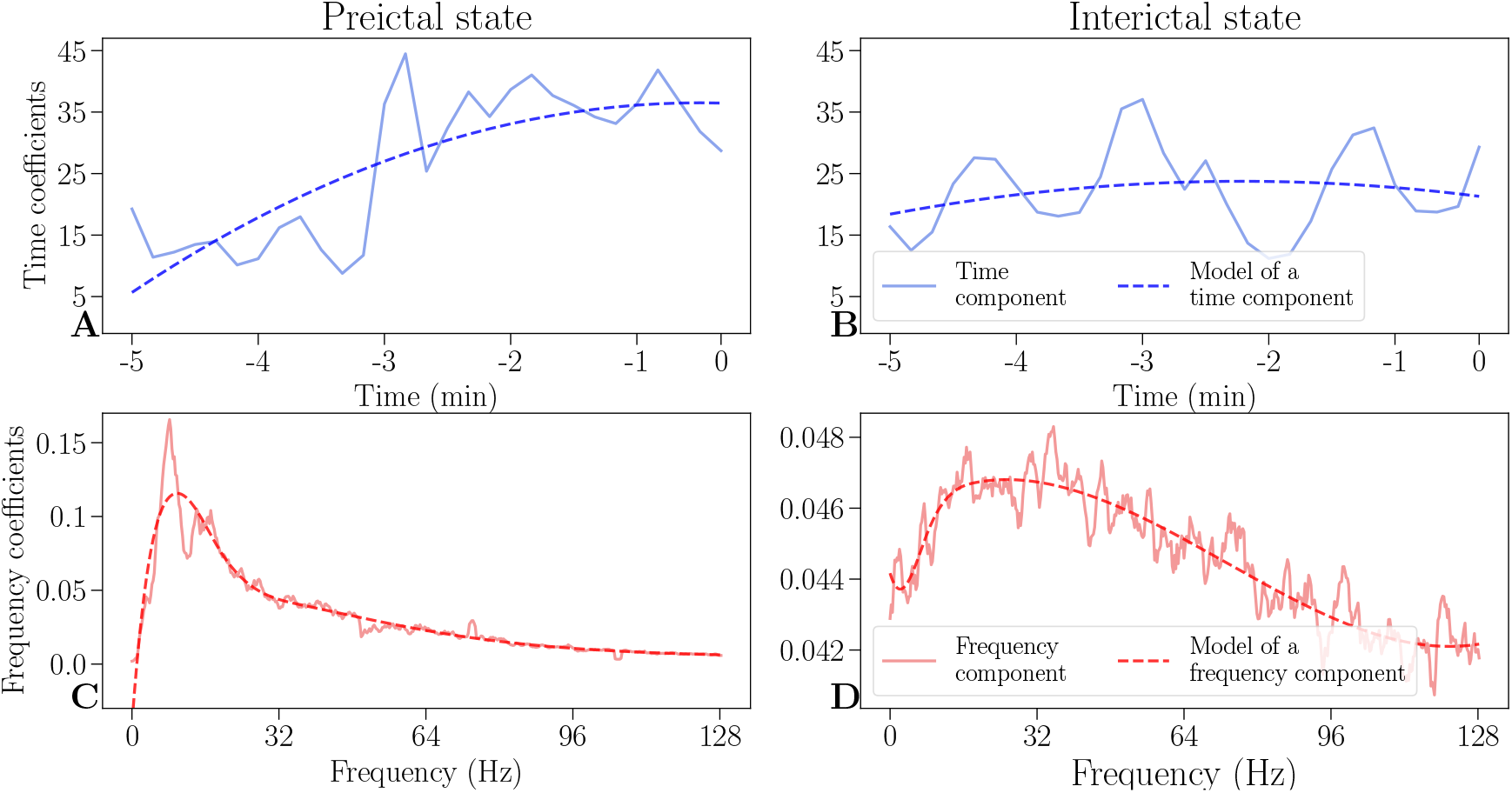
Time and frequency components and its models. An example of decomposed time (solid blue lines) and frequency components (solid red lines) and their respective models (dashed lines) of a preictal state (**A, C**), as well as an interictal state (**B, D**). In a preictal state, the time component (**A**) increases as a seizure is approaching, while the frequency component (**C**) has a sharp increase in low frequencies. Both interictal components (**B, D**) are steady and are an order of magnitude lower than their respective preictal components (**A, C**).

**Figure 3.**
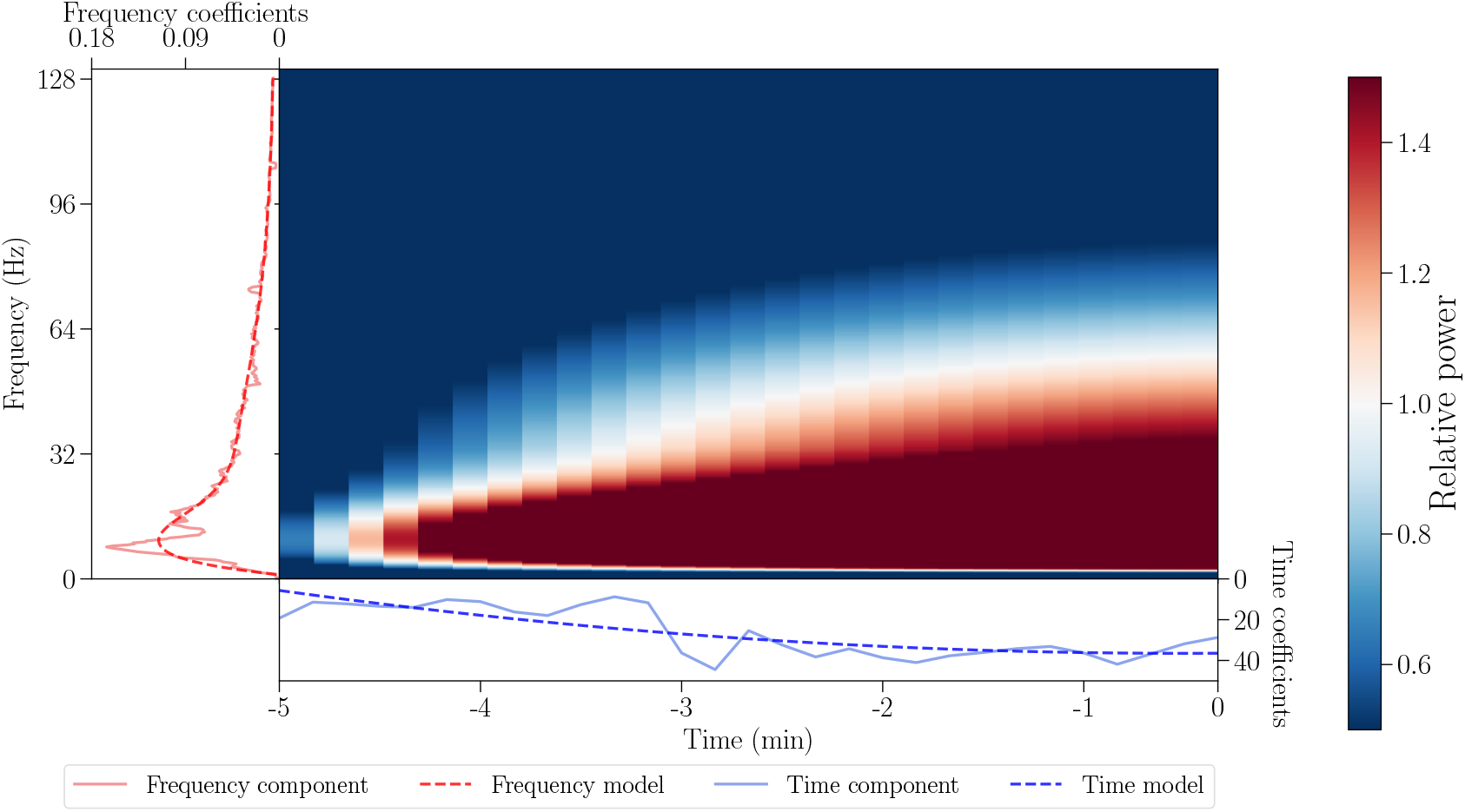
Obtaining a time-frequency model from the respective components. The NMF components are shown with solid red and blue lines for frequency and time, respectively, while their models are shown with dashed lines. The time-frequency model (center) is an outer product of modeled time and frequency components.

### Prediction and performance measures

To classify between preictal and interictal states, linear support vector machines [26] are used. We combine the coefficients of both of the modeled NMF components across all channels into a feature vector. For example, recordings of patient 1 contain 48 channels with 12 NMF parameters (9 parameters for the frequency component and 3 parameters for the time component) each, leading to a dimensionality of 48 · 12 = 576. To account for the risk of overfitting due to the high number of features, L1 regularization is used, which constrains most of the parameters to zero.

In the original dataset, interictal states are more frequent than the preictal ones, which leads to an imbalance of classes (c.f. Tab. 1). To account for this class imbalance, the SMOTE oversampling technique is used, which creates synthetic samples of the minority class based on *k* neighboring points (in our case *k* = 5). [16]

To ensure good generalization of the algorithm, 100-fold cross-validation is used on a training set (70%) and a validation set (30%). Average measures (accuracy, sensitivity, specificity, positive and negative predictive values) are reported. Since the classifier should neither miss nor falsely predict a seizure, we report sensitivity sensitivity and specificity, as well as positive and negative predictive values. [27]

## Results and discussion

### Interpretability of the model

The modeled NMF components show clear differences in preictal and interictal states. [Fig.2] In a preictal state, the modeled time component shows an increase towards the seizure onset, which is not visible in an interictal state. Models of the frequency components of preictal states exhibit a peak of high activity in lower frequencies, relative to baseline activity. Our study thus confirms previous findings of a structure below 30Hz (gamma range), which is informative for seizure prediction. [9, 10] These structural differences are also visible in recovered time-frequency models (see S2 Figure and S3 Figure in the appendix). Even though there are differences between patients in both time and frequency components, the average components are similar among patients, which confirms the reliable existence of structure in preictal states (see S4 Figure in the appendix for a comparison).

### Predictive performance

Similar accuracy is achieved for all patients (above 90%). The lowest performance is for the patient 5 (92%) and the highest for the patient 4 (100%), as shown in figure 4 and table 2. Sensitivity is between 0.84 and 1, while specificity ranges from 0.98 to 1, as can be seen in figure 4. A combination of high values of sensitivity and specificity is achieved for all patients. Similarly, positive predictive values are between 0.98 and 1, while negative predictive values are between 0.87 and 1 (c.f. figure 4 and table 2).

**Table 2.**
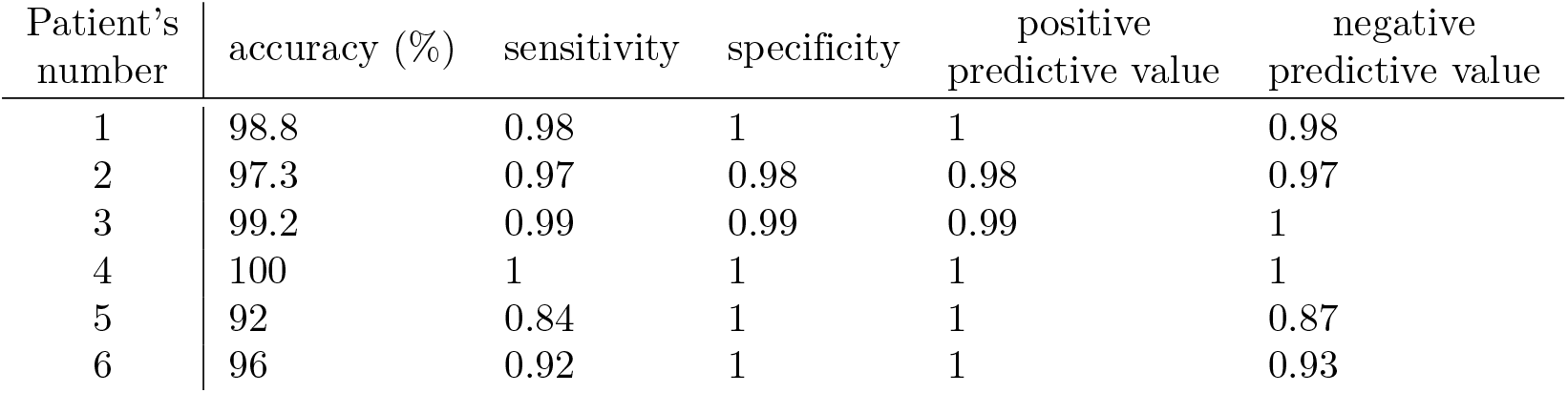
Performance measures for all patients.

**Figure 4.**
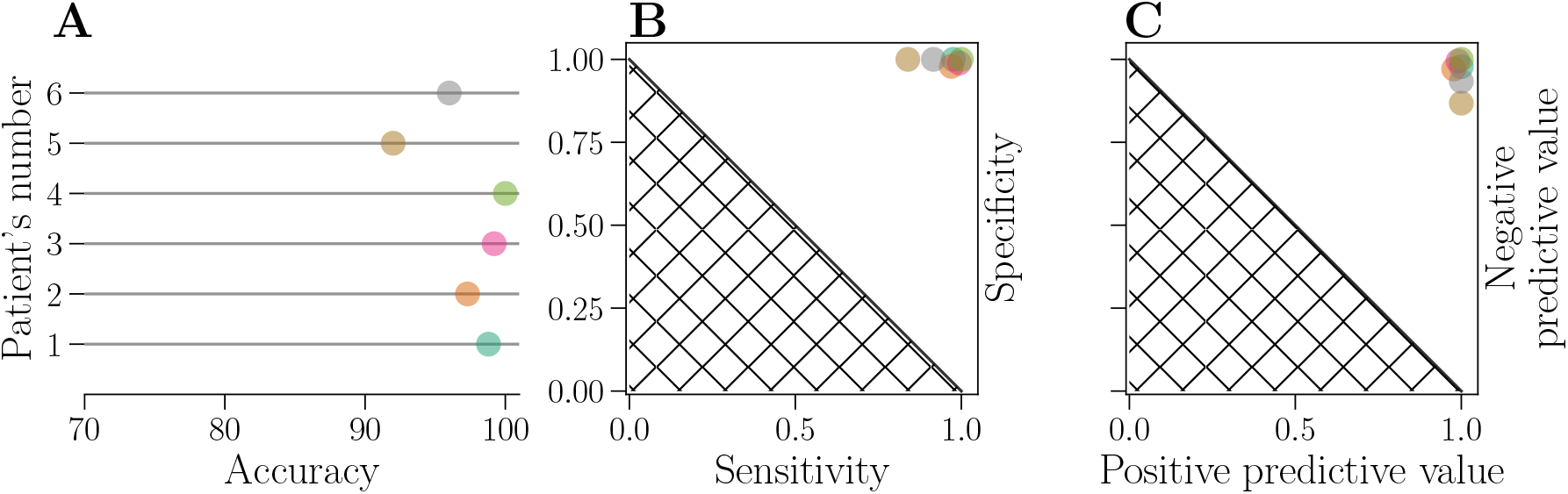
Evaluation of prediction performance. Performance of each patient is represented by a circle, for accuracy (**A**), specificity-sensitivity plot (**B**) and negative and positive predictive value (**C**). Identical colors are used to represent each patient across all three subplots. The hatched area represents results attainable by a random classifier.

## Conclusion

Since patients with uncontrolled epilepsy prefer to be advised a few minutes before a seizure onset [18], we decided to use intervals of five minutes. However, this method is easily extensible to longer periods of time, since the length of intervals has no effect on dimensionality of modeled time components.

Data from additional patients as well as more data from the same patient could, if available, lead to a better generalization of the model. This however is a challenge for patient-specific models in general, where data from a single patient should suffice, and a large number of labeled training examples is not available.

Overall, this study demonstrates the use of nonnegative matrix factorization of power spectra for a seizure prediction task. The proposed model is conceptually simple, interpretable and has shown high accuracy on a representative dataset. A similar approach could be used for similar tasks such as detection of sleep stages in EEG or the detection of irregularities in ECG.

## Data Availability

The data used in this project is a part of the EPILEPSIAE dataset, which is not publicly available. For this reason, there are no specific paths or identification numbers in the code repository which is publicly available.
The code is provided here: https://github.com/ostojanovic/seizure_prediction

## Supporting Information

**S1 Figure.**
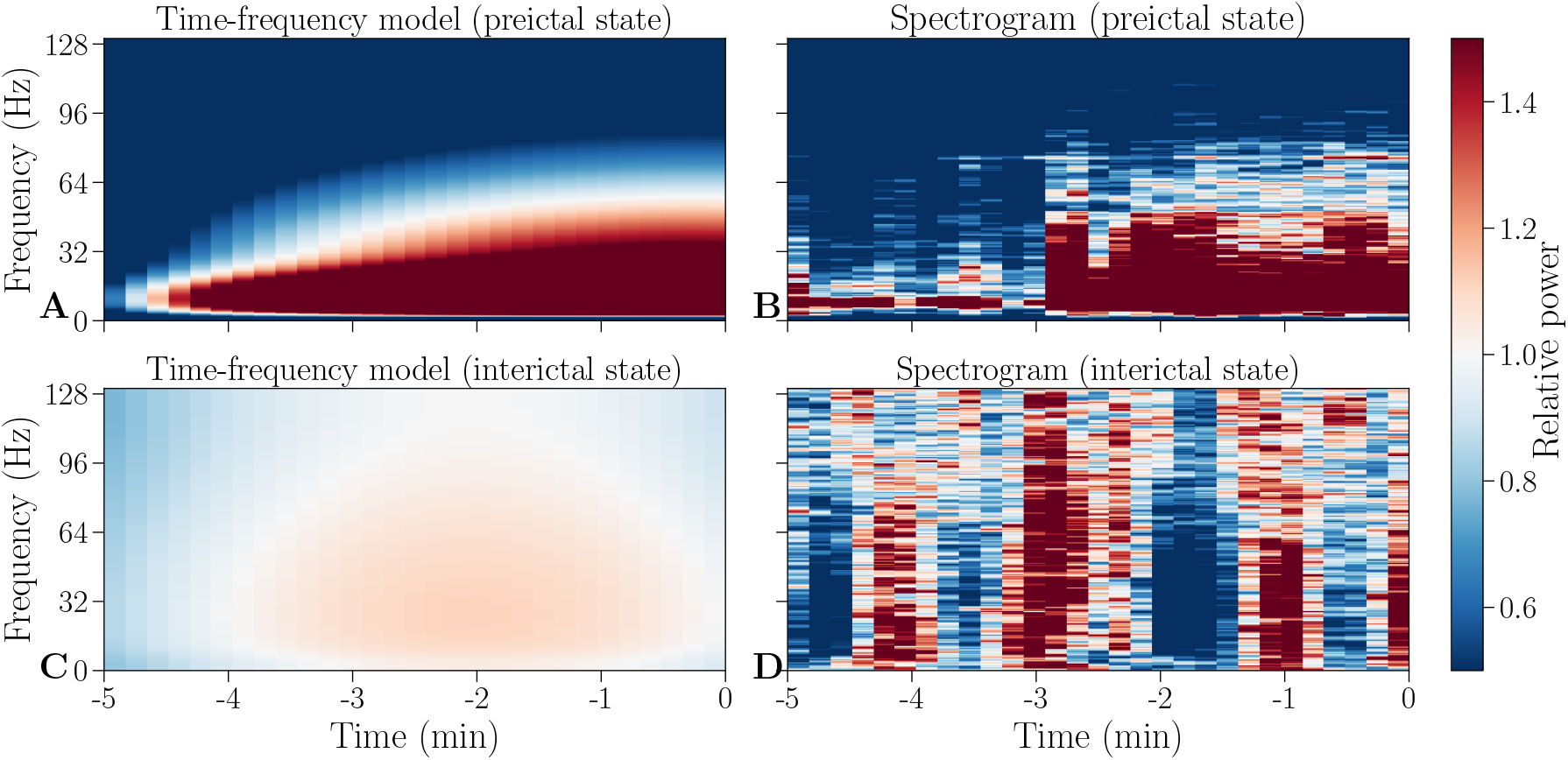
Time-frequency models and corresponding spectrograms of preictal and interictal states. An outer product of modeled time and frequency components (**A, C**) and corresponding spectrograms (**B, D**). A preictal state is shown in the upper row (**A-B**) and an interictal state is shown in the bottom row (**C-D**).

**S2 Figure.**
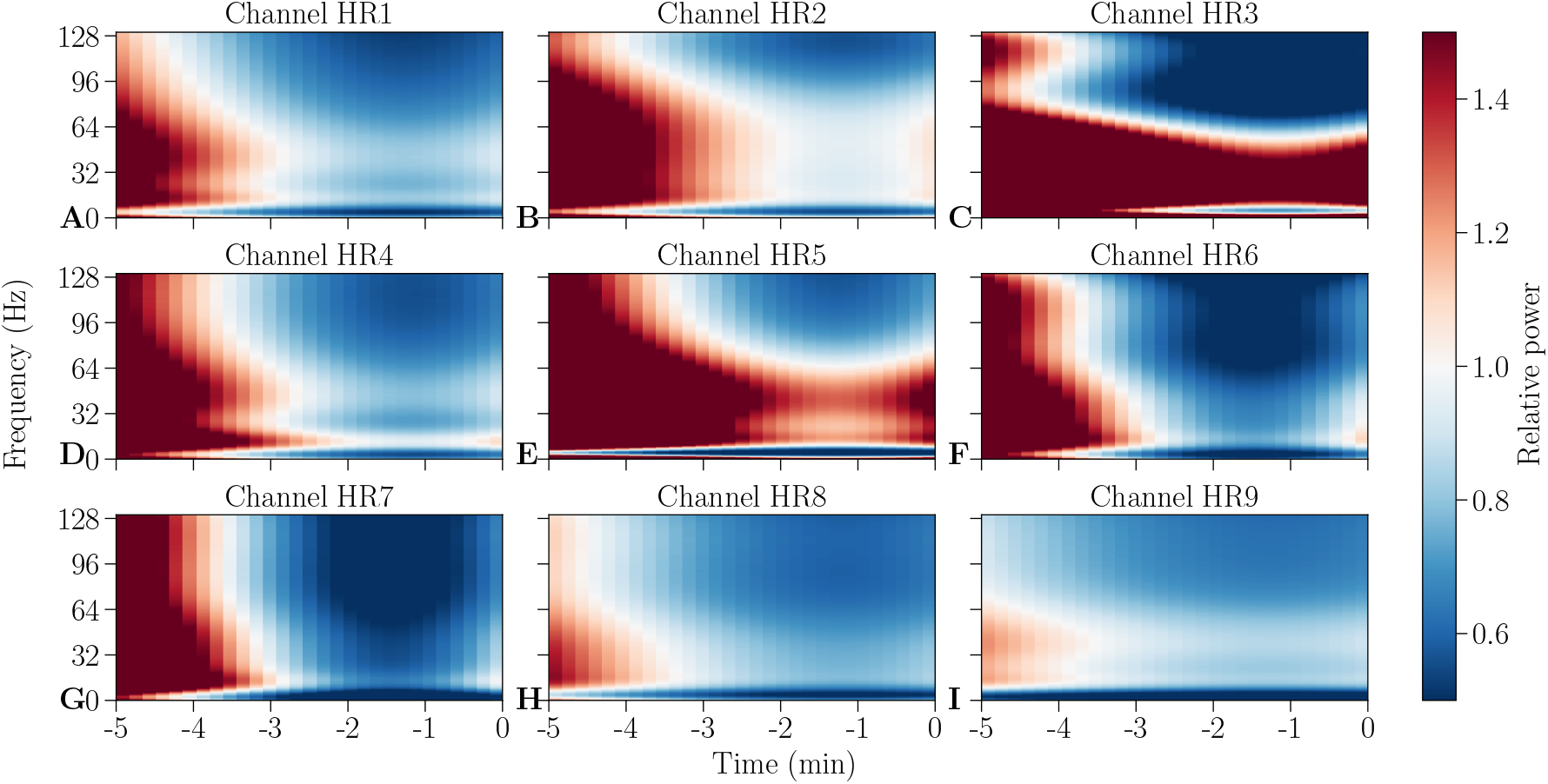
Models of preictal states. Models shown here are for different channels (**A-I**) from the same individual measurement period for patient 1.

**S3 Figure.**
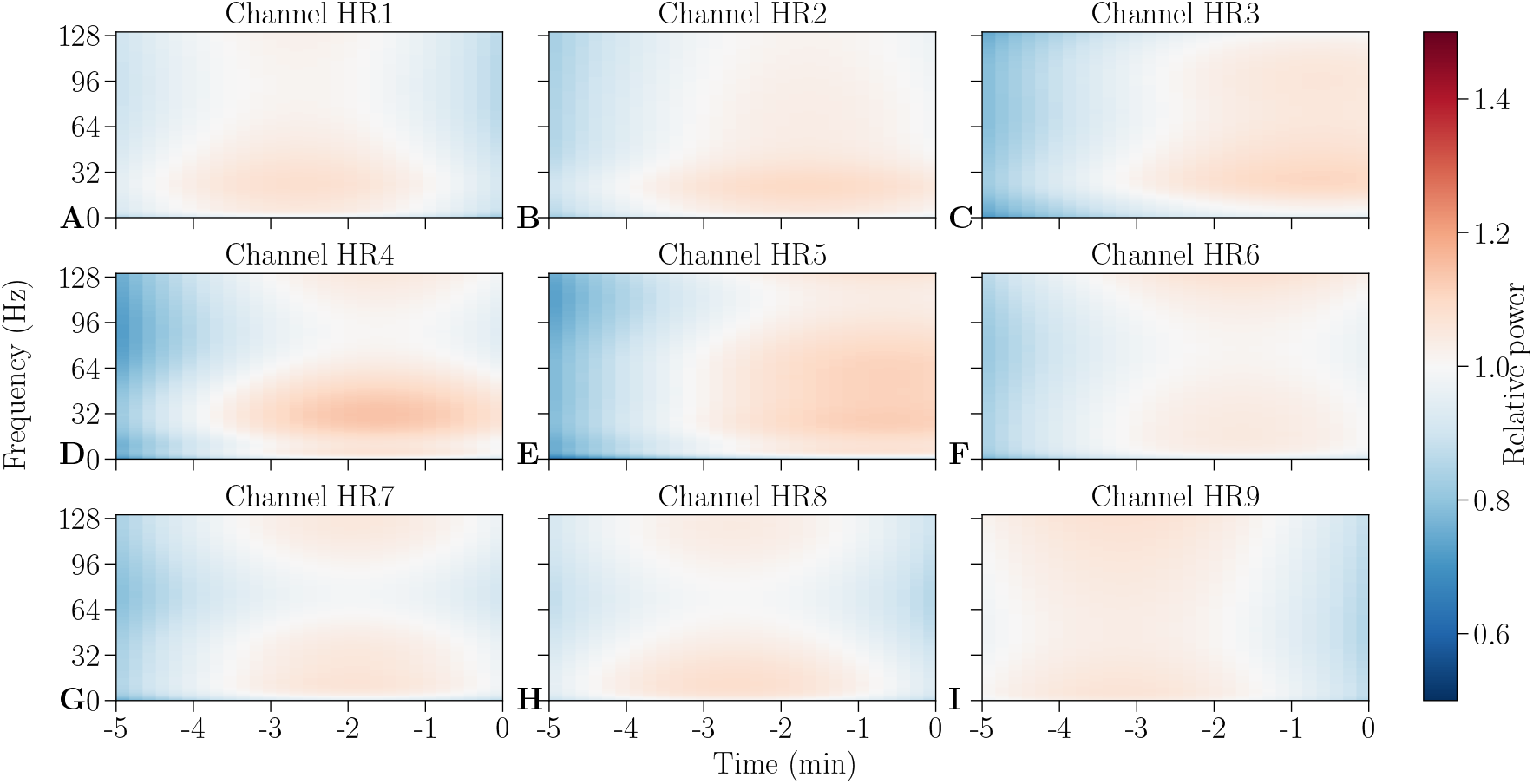
Models of interictal states. Models shown here are for different channels (**A-I**) from the same individual measurement period for patient 1

**S4 Figure.**
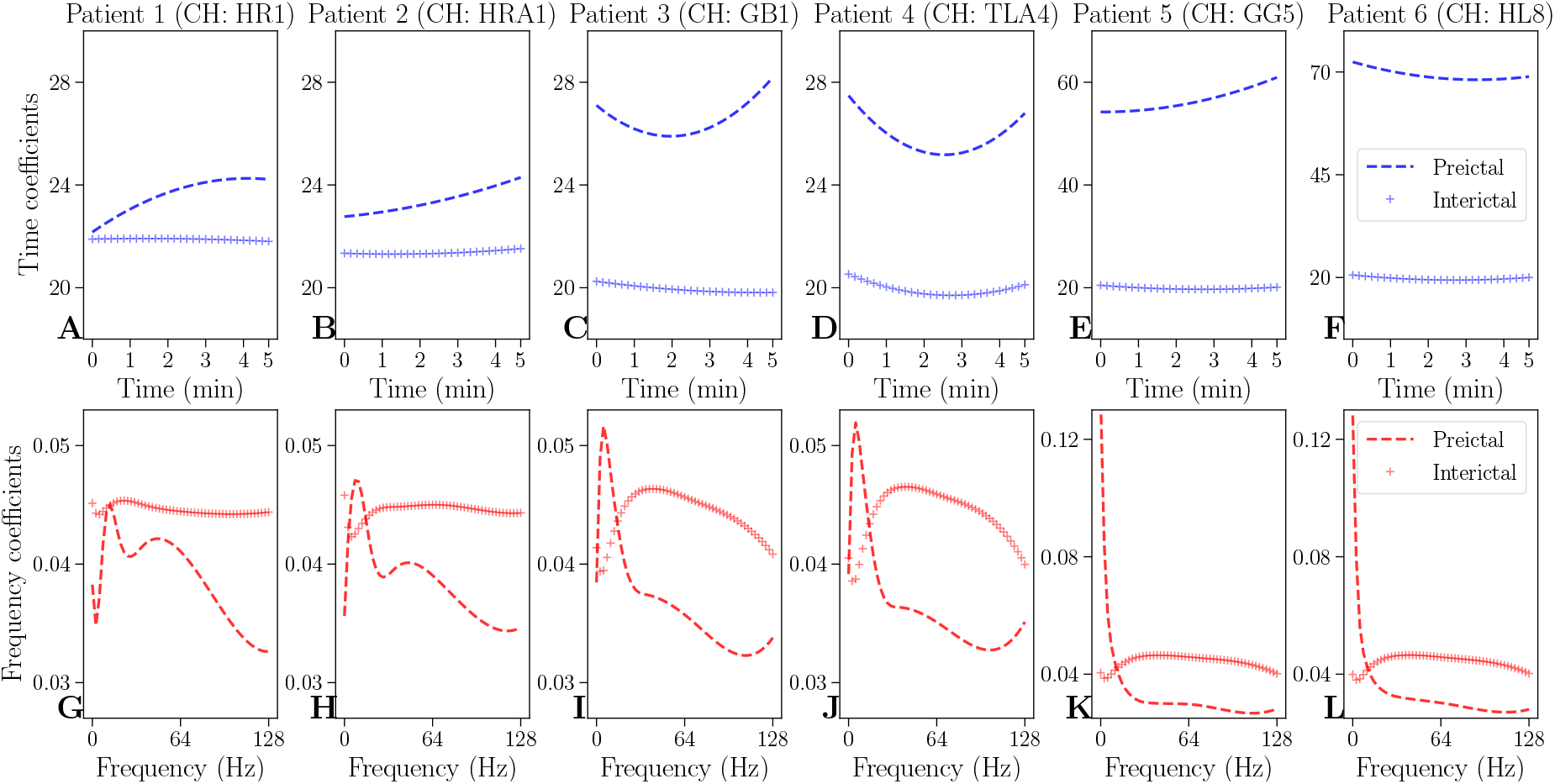
Models of time and frequency components for one measurement and one electrode for preictal and interictal states for all six patients. Models of time components are shown in the upper row (**A-I**), and models of frequency components are shown in the bottom row (**G-L**). Preictal states are indicated with a dashed line and interictal states are indicated with a line marked with + in blue for models of time and red for models of frequency components, respectively.

## Acknowledgments

We are very grateful for early discussions with the team of the Freiburg Epilepsy Center, especially Prof.Dr.med. Andreas Schulze-Bonhage and Dr.-Ing. Matthias Dümpelmann, and to helpful input from our colleague Johannes Leugering.

## Footnotes

1 A software implementation of the presented method is available online at https://github.com/ostojanovic/seizure_prediction.

